# Forecasting daily COVID-19 confirmed, deaths and recovered cases using univariate time series models: A case of Pakistan study

**DOI:** 10.1101/2020.09.20.20198150

**Authors:** Hasnain Iftikhar, Moeeba Rind

## Abstract

The increasing confirmed cases and death counts of Coronavirus disease 2019 (COVID-19) in Pakistan has disturbed not only the health sector, but also all other sectors of the country. For precise policy making, accurate and efficient forecasts of confirmed cases and death counts are important. In this work, we used five different univariate time series models including; Autoregressive (AR), Moving Average (MA), Autoregressive Moving Average (ARMA), Nonparametric Autoregressive (NPAR) and Simple Exponential Smoothing (SES) models for forecasting confirmed, death and recovered cases. These models were applied to Pakistan COVID-19 data, covering the period from 10, March to 3, July 2020. To evaluate models accuracy, computed two standard mean errors such as Mean Absolute Error (MAE) and Root Mean Square Error (RMSE). The findings show that the time series models are useful in predicting COVID-19 confirmed, deaths and recovered cases. Furthermore, MA model outperformed the rest of all models for confirmed and deaths counts prediction, while ARMA is second best model. The SES model seems superior to other models for prediction of recovered counts, however MA is competitive. On the basis of best selected models, we forecast form 4*th* July to 14*th* August, 2020, which will be helpful for decision making of public health and other sectors of Pakistan.

## 1 Introduction

COVID-19 is an infectious disease, which grows rapidly in populous areas. The World Health Organization (WHO) declared COVID-19 as a world-wide pandemic has appeared as the most destructive disease impacting at least 99% countries of the world and first identified in Wuhan City, Hubei Province, China [1]. The humanitarian costs of the COVID-19 outbreak have been rising since 31*st* December, 2019 as it affected more than 10,710,005 people and deaths counts 517,877 were till 03 July, 2020 globally [2]. The countries with Pakistan’s borders infected by COVID-19, including Iran and China, which was the major cause of effecting Pakistani’s. The first two cases confirmed on the 26*th* February, 2020, in Islamabad and Karachi [3]. Due to a weak health system of the country, many peoples are effected and careless public attitude and mega shopping made the coming days worst. On the 13*th* March, the Government of Pakistan has imposed complete lack-down in the whole country and took the initial steps for reducing the spread of virus; cancelled conferences to disrupted supply chains, imposed travel restrictions, closing of borders, tremendously wedged travel industry, close flights and within country disrupted work, closing of shopping mall, school, colleges and universities. For awareness of peoples different TV programs, commercial and advertisements were organized. Face mask and sensitizer were used by each and every person[4].

Since the mildness in lockdown on April 15, 2020 and then after further relaxation since 12 May 2020 the number of cases increased manifold. During remaining days of May more than fifty thousand new cases added. The rise did not stop there. The month of June proved to be worse. AS, the total number of confirmed and deaths counts in the country till 3*rd* July, 2020 were 198,883 and 4,035 respectively. Sindh has reported the highest cases that are 76,318, followed by Punjab with 72,880 cases whilst Punjab has recorded the highest deaths in a country, a total of 1,656 followed by Sindh with 1,205 deaths [5]. A continuous struggle is required to occupy the spread of COVID-19 in such a way that health sector can deal with COVID-19 patients in the future.

Currently, several studies have been undertaken to predict the behaviour of virus [6–12]. For example, [13] used Autoregressive Integrated Moving Average model (ARIMA) in order to predict number of COVID-19 deaths and recoveries for Pakistan. The work in [14] proposed three phase Susceptible-Infected-Recovered-Dead (3P-SIRD) model compute a supreme lock-down period for several particular geographical areas to break the transmission chain of virus and help country to recover. The authors [15] in, forecast the epidemic peak eruption of the COVID-19 in Turkey, Brazil and South Africa using age structured SEIR system. Some researchers predicted the continuation of the COVID-19 using exponential smoothing method. For example, [16] explored the development of informational efficacy in crypto-currency markets as well as international stock markets before and during the pandemic caused by COVID-19. They found that cryptos are more in-stable during the novel COVID-19 pandemic than international stock markets. Thus, making investment in digital assets during the pandemic times might be riskier. Few authors used machine learning models for forecasting of COVID-19 [17–19]. In the work [20] investigated that, the spread of COVID-19 using the case of Malaysia and scrutinized its linkage with some external factors e.g. inadequate medical resources and incorrect diagnosis problems. They have used epidemiological model and dynamical systems technique and observed that might misrepresent the evaluation on the severity of COVID-19 under complexities. In order to forecast agreement to the publicly available data, the work in [21] used Fractional time delay dynamic system (FTDD). The author in [22] used Generalized logistic model and found the pandemic growth as exponential in nature in China. The author in [23] used genetic programming (GP) models for confirmed cases and death cases in three highly COVID-19 affected states of India i.e. Maharashtra, Gujarat, Delhi and whole India. They have statistical validated the evolved models to find that the proposed models based on GEP use simple interactive functions and can be highly relied upon time series forecasting of COVID-19 cases in the context of India. Based on the spreading behaviour of the COVID-19 in the mass, [24] estimated three novel quarantine epidemic models. They found that isolation at home and quarantine in hospitals are the two most effective control strategies under the current circumstances when the disease has no known available treatment. In the work [25] using positive cases over 50 days of disease progression for Pakistan, analysed the graphical trend and using exponential growth forecasted the behaviour of disease progression for next 30 days. They assume different possible trajectories and projected estimated 20k-456k positive case within 80 days of disease spread in Pakistan.

Due to the mutated nature of the virus, the situation has become graver with little known about the cure, there remain greater uncertainty about the probable time-line of this disease. Hence, forecasting for short term is immensely important to get the clue for predicting the flattening of curve and revival of routine social and economic life [26]. Statistical models using evidence from real world data can help predict the location, timing, and the size of outbreaks, allowing governments to allocate resources more effectively, to conduct scenario and signal analysis, and to determine policy approaches. Epidemiological tools can then be applied to limit the scope and spread of outbreaks. However, these approaches are sensitive to the underlying assumptions and hence impact vary [27]. It is important to ensure oversight, check assumptions in modelling; and ensure the veracity, reliability, and accountability of these tools in order address bias and other potential harms. In this work, attempt to look at the projections for COVID19 infections of Pakistan, using a number different univariate time series methods.

The rest of article is arranged as: Section two described forecasting models and three disused the out-of-sample and forecasting results. Finally, Section four comprises of conclusion and discussion.

## 2 Forecasting Models

In this work, we consider five different univariate time series models including; Autoregressive (AR), Moving Average (MA), Autoregressive Moving Average (ARMA), Nonparametric AutoRegressive (NPAR) and Simple Exponential Smoothing (SES). These models are described with detail in the following:

### 2.1 Autoregressive Process

A linear Autoregressive (AR) process describes a linear function of the previous n observations of *M*(*t*), is defined as:

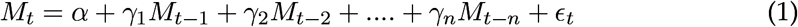

where *α* and *γ*_*i*_(*i* = 1, 2, …, *n*) are the intercept and slope coefficients of the underlying AR process and *ϵ*_*t*_ is the disturbance term. After, an examination graphical analysis (plotting the series residuals, ACF and PACF), fit an AR(2) *M*_*t*_ to each time series.

### 2.2 Moving Average Model

Moving Average (MA) model is primarily remove the periodic fluctuations in the time series data, for example fluctuations due to seasonality. The Moving average model mathematically can be written as:

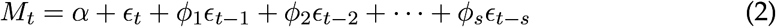

*α* indicate the constant (intercept), *ϵ*_*j*_ (*j* = 1, 2, …, *s*) are parameters of MA model and the *ϵ*_*j*_ is white process. The values of s are revealing the order of the MA process.

### 2.3 NonParametric Autoregressive Model

The additive nonparametric counterpart of AR process leads to additive model, where the association between *M*_*t*_, and its previous lags have non-liner relationship, which may be describe as:

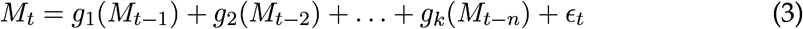

where *g*_*i*_ are showing smoothing functions and describe the association between *M*_*t*_ and its previous values. In the recent case, functions *g*_*i*_ are denoted by cubic regression splines. As in case of parametric form, we utilized 2 lags while estimating NPAR.

### 2.4 Autoregressive Moving Average Model

Autoregressive Moving Average (ARMA) model can be define as, the response variable *M*_*t*_ is regressed on the previous n lags also with residuals (errors) as well. Mathematically,

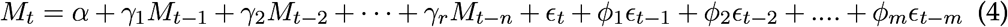

where *α* denotes intercept, *γ*_*i*_(*i* = 1, 2, …, *n*) and *ϕ*_*k*_(*k* = 1, 2,, *m*) are the parameters of AR and MA process respectively, and *ϵ*_*t*_ is a Gaussian white noise series with mean zero and variance 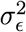. The ARMA model order selection is established through inspecting the correlograms (i.e. Partial and Auto-correlation function (P-ACF)). In our case, fit an ARMA (1, 1) model to each series *M*_*t*_.

### 2.5 Simple Exponential Smoothing Model

The Simple exponential smoothing (SES) model of forecasting allows the researchers to smooth the time series data and then use it for out of sample forecasting. SES model is applicable when the data is stationary i.e., no trend and no seasonal pattern but the data at level changing gradually over time.

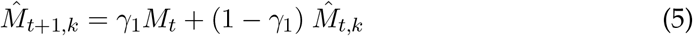

where *γ*_1_ is the smoothing constant, *M*_*t*_ is showing the actual series, 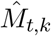 is representing the forecasted value of the underlying series for period *t* and 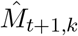 is denoting the forecasted value for the period *t* + 1. This method assigns the weights in such a way that moving back from the recent value, the weights exponentially decreases. For the modelling purpose, a prime assumption of time series data is stationarity. A stationary process is defined as that the mean, variance and autocorrelation structure are time invariant. If the underlying series is nonstationary, it must be transform to stationary. In the literature, different techniques are used to achieve stationarity, for example, taking natural log, differencing the series or box-cox transformation etc [28]. In this work, the COVID-19 confirmed, deaths and recovered counts times series are plotted in Figure 1 **(left-column)** daily and Figure 1 **(right-column)** cumulative cases. Clearly seen, all the three daily time series having an upward increasing linear trend, which show that the series is non-stationery, hence need to make stationary using differencing method. Also, to check the unit root issue of the underlying series that are conformed, deaths and recovered cases, we apply Augmented Dickey Fuller test (ADF) test. The results are tabulated in Table1, which suggested that the all three series are non-stationary at level. However, taking first order difference, the series are turned out to be stationary. The first order differencing series of daily confirmed, deaths and recovered cases are piloted in Figure 2, where now the series do not contain any trend, hence its become stationary.

**Figure 1:**
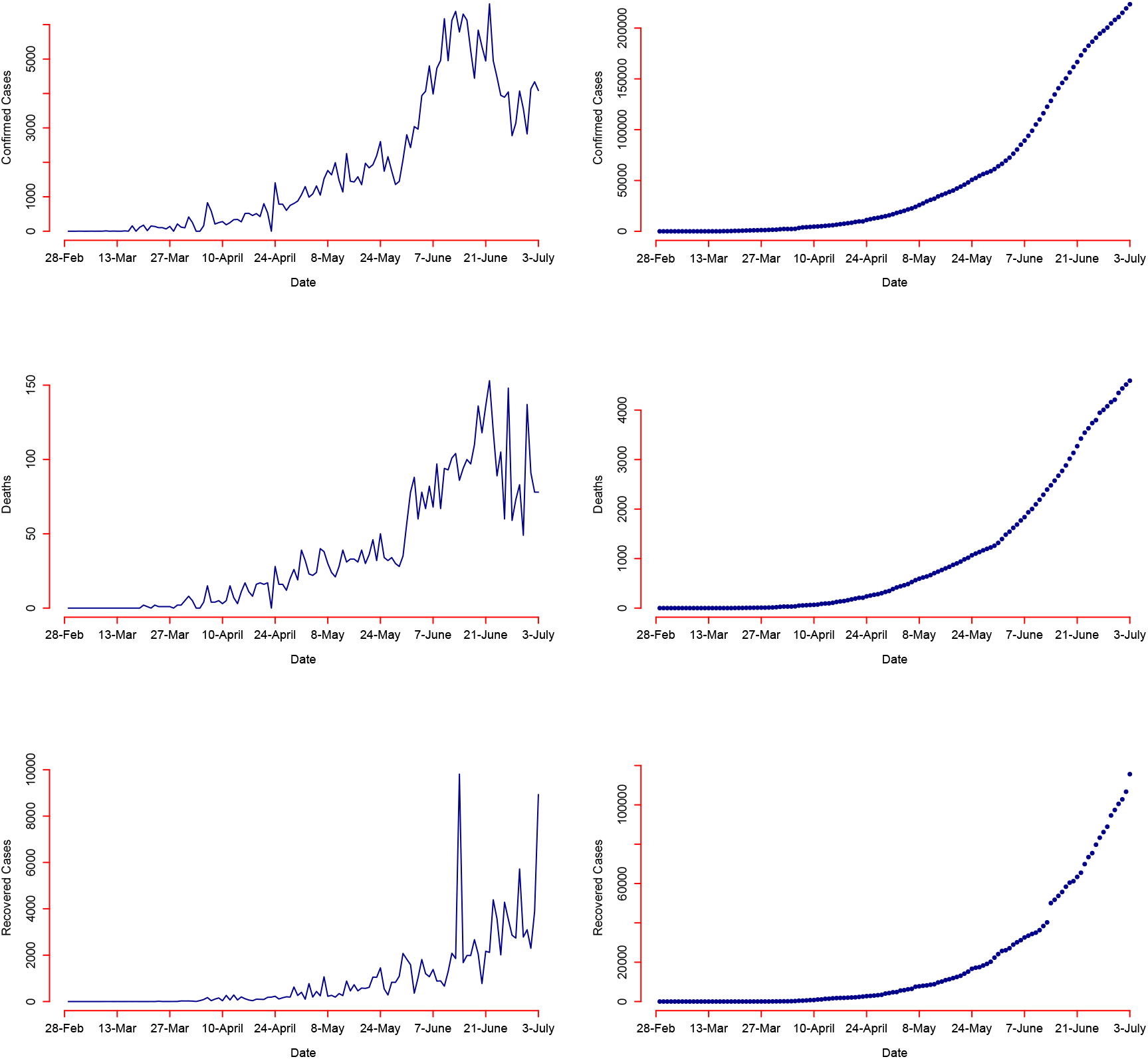
Pakistan COVID-19: Confirmed, deaths and recovered counts daily (**left-column**), and cumulative (**right-column**) over the period of 28, February to 03, July 2020.

**Figure 2:**
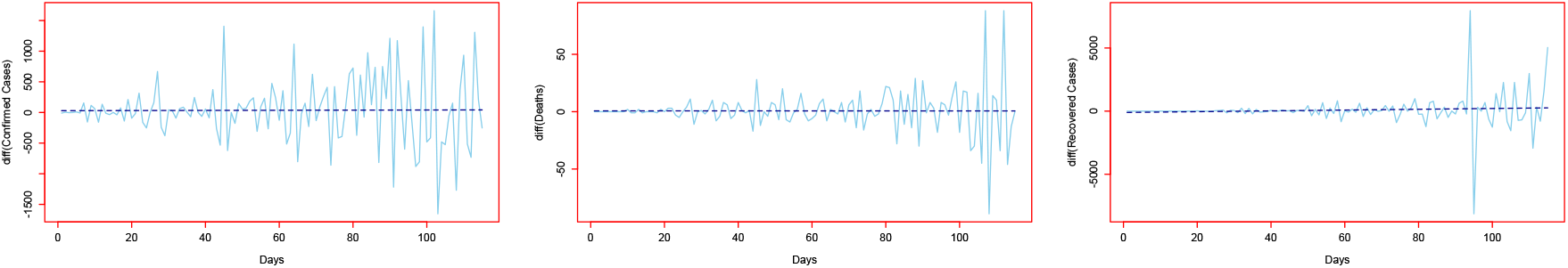
Differenced series: 1st order difference for Confirmed (**left**), Deaths (**middle**) and Recovered cases (**right**).

## 3 Results

In this paper, we used daily COVID-19 conformed, deaths, and recovered cases for Pakistan. The dataset was obtained by WHO[3], the each series ranges from 10, March 2020 to 3, July 2020. The complete dataset covers 116 days, of which data from 10, March 2020 to 19, May 2020 (71 days) were used for model training and from 21, May to 3, July 2020 (45 days) for one-day ahead post-sample (testing) predictions. For the predicting accuracy, two accuracy measures, Root Mean Square Error (RMSE) and Mean Absolute Error (MAE) for each model were computed as follows:

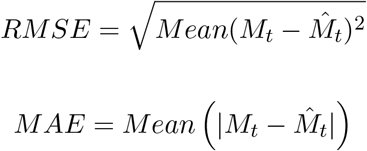

where *M*_*t*_ = Observed and 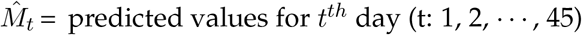.

To evaluate the best model of among the previously described models for each series, we computed two standard accuracy measures and presented the outcomes in Table 2 to 3. From the output in both Tables 2 and 3, we can observe that MA model produced low errors as compare to all other competitors for the confirmed and deaths counts predications. The RMSE and MAE values for MA model are 733.92 and 629.95 for conformed and 24.78 and 18.02 for deaths counts, respectively. However, ARMA model is competitor. The prediction of recovered patient of COVID-19, SES model shows better results as compared to rest of all models, while MA model is second best model. The RMSE and MAE values for SES model are 1897.32 and 1057.09, separately. The RMSE and MAE values for each series that are computed using different five models are also plotted in Figure 3 where the superiority of MA (confirmed and deaths cases)and SES (recovered cases) models can be evidently seen in both cases training and testing exercise.

**Table 1:** Augmented Dickey-Fuller (ADF) test Statistics.

|  | At level | At first difference |  |
| --- | --- | --- | --- |
| Variables | Constant with trend | Constant with trend | Conclusion |
| Cases | -1.806 | -10.447* | I (1) |
| Deaths | -1.022 | -7.470* | I (1) |
| Recoveries | -0.095 | -6.348* | I (1) |

**Table 2:** Model Estimation/Train: One-day-ahead RMSE and MAE for confirmed, deaths, and recovered cases for all models.

| Model Estimation/Train |  |  |  |  |  |  |
| --- | --- | --- | --- | --- | --- | --- |
| MODELS | Conformed |  | Deaths |  | Recovires |  |
|  | RMSE | MAE | RMSE | MAE | RMSE | MAE |
| AR | 385.02 | 268.58 | 9.48 | 6.65 | 647.34 | 338.87 |
| MA | **371.74** | **252.98** | **9.21** | **6.00** | 574.14 | 220.60 |
| NPAR | 393.85 | 277.07 | 9.26 | 6.42 | 564.36 | 264.02 |
| ARMA | 380.69 | 256.66 | 9.31 | 6.06 | 552.89 | 247.70 |
| SES | 383.17 | 257.93 | 9.42 | 6.03 | **544.18** | **206.87** |

**Table 3:** Out-of-Sample/Test: One-day-ahead RMSE and MAE for confirmed, deaths, and recovered cases for all models.

| Out-of-Sample/Test |  |  |  |  |  |  |
| --- | --- | --- | --- | --- | --- | --- |
| MODELS | Conformed |  | Deaths |  | Recovires |  |
|  | RMSE | MAE | RMSE | MAE | RMSE | MAE |
| AR | 755.07 | 620.95 | 25.65 | 19.17 | 2500.20 | 1349.63 |
| MA | **733.92** | **629.95** | **24.78** | **18.02** | 1987.75 | 1059.44 |
| NPAR | 824.53 | 711.87 | 33.39 | 24.79 | 2623.00 | 1264.31 |
| ARMA | 743.24 | 636.31 | 25.46 | 19.36 | 2143.37 | 1173.68 |
| SES | 782.89 | 661.09 | 25.60 | 18.55 | **1897.32** | **1057.09** |

**Figure 3:**
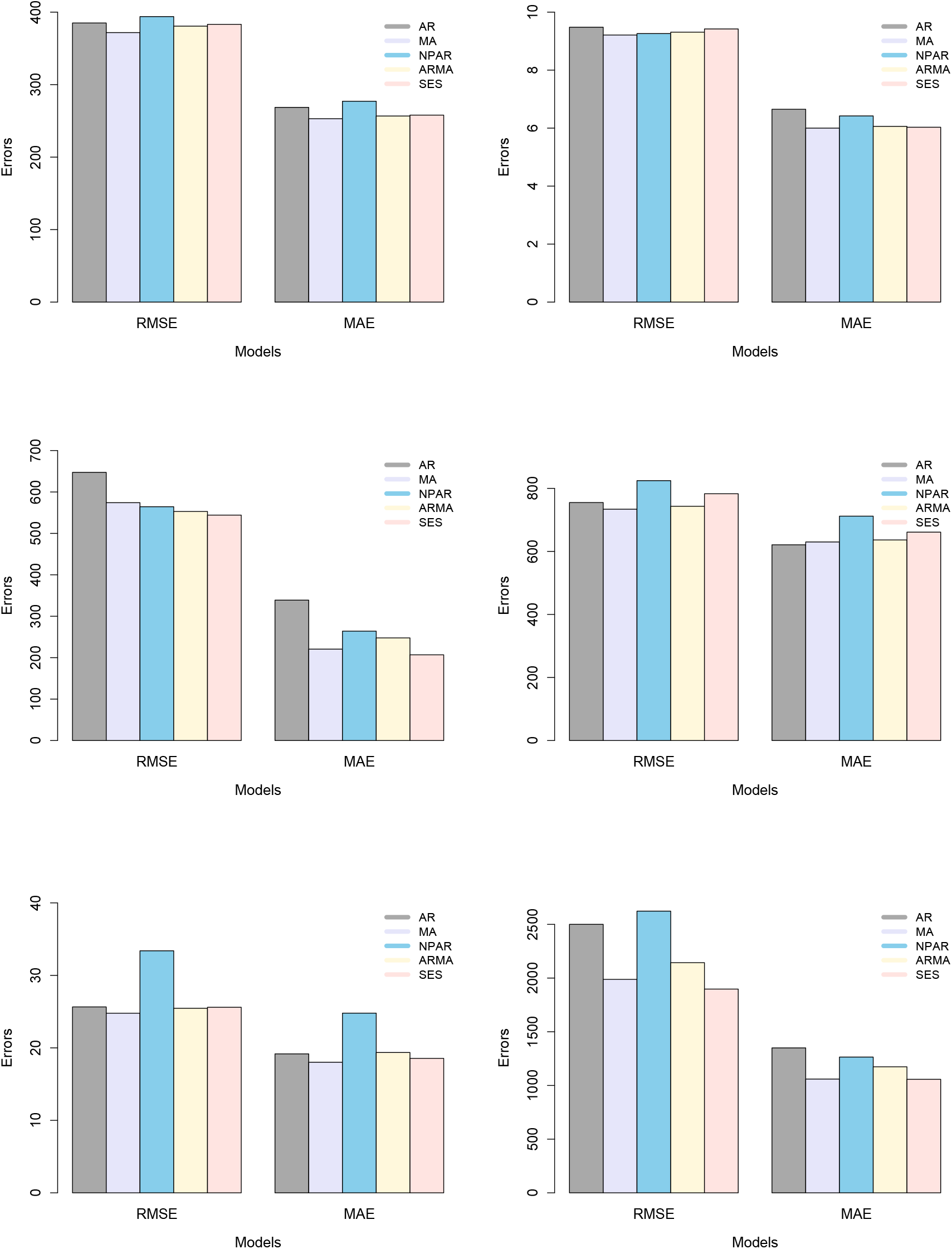
Barplot: RMSE and MAE for confirmed, deaths and recovered cases; **Model Estimation/Train (left-column), Out-of-Sample/Test (2nd-column)** for all models.

The day-specific confirmed, deaths and recovered case are plotted in Figure 4, over the period of 21, March to 19, June 2020. From the Figure 4(left-column) can be observed that variation among the different weeks, while Figure 4(right-column) mean of days are plotted for conformed, deaths, and recovered cases. where clearly seen that the an increasing pattern Saturday to Friday, which is show that the effect of working and non-working days.

**Figure 4:**
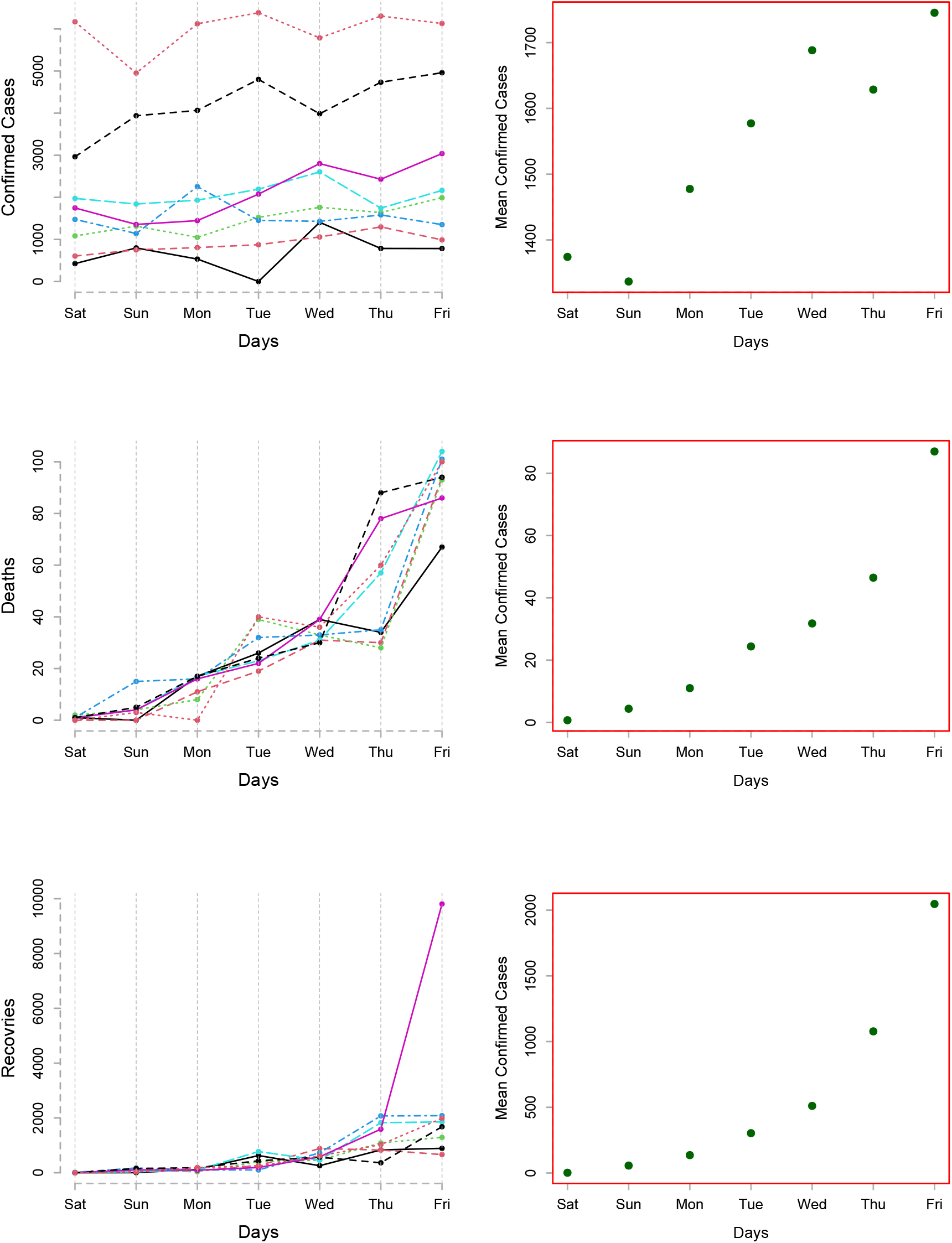
Weekly COVID-19 Cases: Day-specific confirmed, deaths and recovered cases; **(left-column)** and mean day-specific **(right-column)**for the period of 21, March to 19, June 2020.

Once the best models assessed through the out-of-sample mean errors (RMSE, MAE), then we proceed for future forecasting with the superior model in each case. We used MA for confirmed and deaths cases and SES for recovered cases and forecast from 4, July to 14, August 2020 for both daily and cumulative cases. The forecasted values are seen in Figures 5, clearly revealing that deaths and recovered cases are monotonically increasing, while conformed counts are not. The confirmed cases on 14, August 2020 are expected 7,325 and cumulative cases 413,639, deaths during the end of mid August are expected 121 and cumulative counts are 9,279, and the recovered cases are 10,730 and cumulative are 455,661. Overall, the results suggested that, the increasing of confirmed case are gradually decreased, which was the outcome of Government imposed earlier steps such as cancelled conferences to disrupted supply chains, imposed travel restrictions, closing of borders, tremendously wedged travel industry, close flights and within country disrupted work, closing of shopping mall, school, colleges and universities. For awareness of peoples different TV programs, commercial and advertisements were organized. Face mask and sensitizer were used by each and every person.

**Figure 5:**
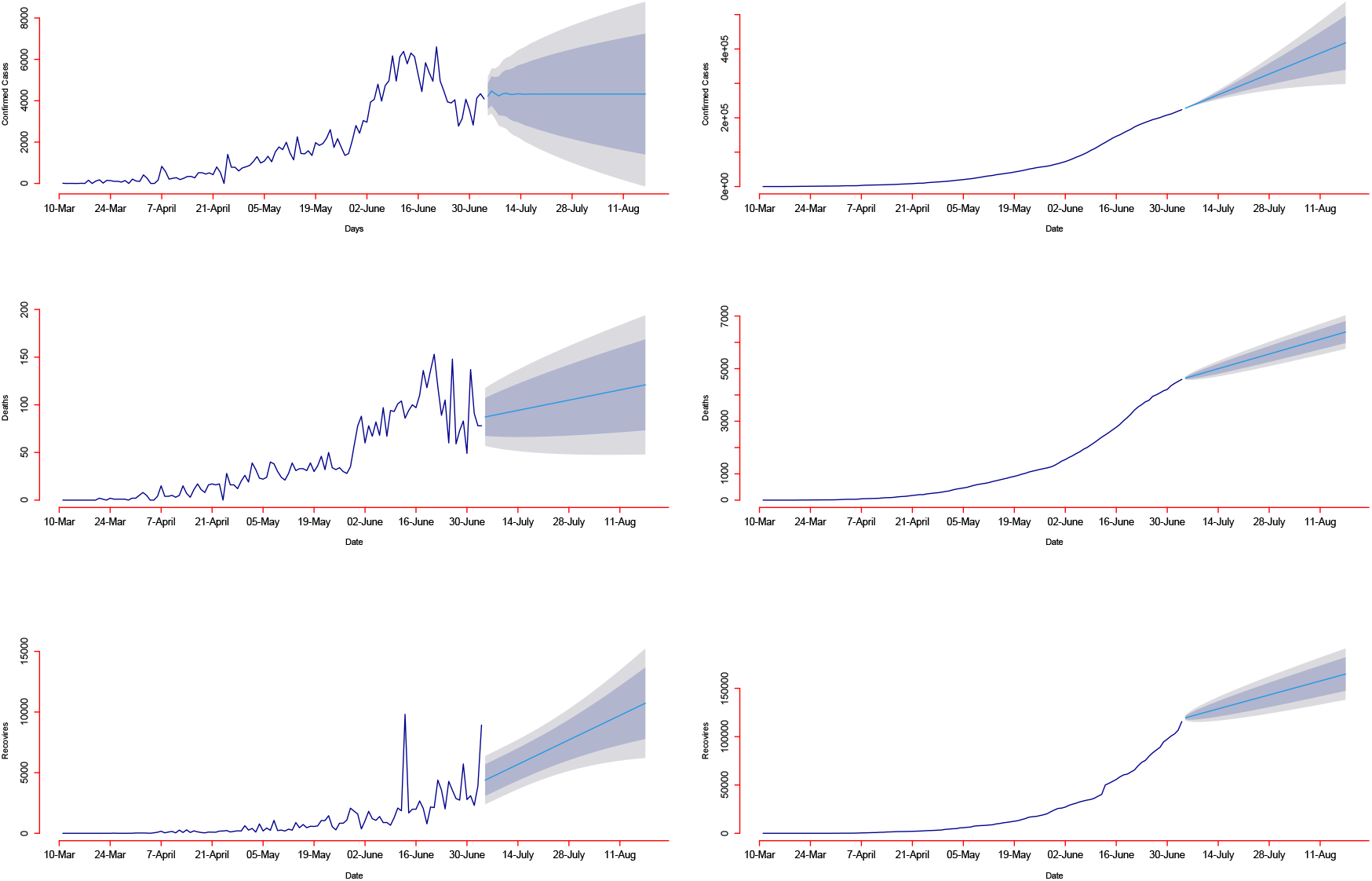
Forecasts COVID-19: Confirmed daily and cumulative cases by MA Model **(1st row)**, Deaths daily and cumulative cases by AR Model **(2nd row)** and Recovered daily and cumulative cases By SES Model **(3rd row)** for the period 3, July to 14, August 2020.

## 4 Conclusion

The main purpose of this work was to forecast confirmed, deaths and recovered cases of COVID-19 for Pakistan using five different univariate time series models including; Autoregressive (AR), Moving Average (MA), Autoregressive Moving Average (ARMA), Nonparametric Autoregressive (NPAR) and Simple exponential smoothing (SES) models. The dataset of confirmed, deaths and recovered cases ranges from 10, March to 03, July 2020 was used. For model estimation/training was used from 10, March 2020 to 19, May 2020 and 20, May to 3, July 2020 were used for one-day-ahead out-of-sample predictions. To check the predicting performance of all models, we use RMSE and MAE as mean errors. Moreover, MA model beat the rest of all models for confirmed and deaths counts prediction and SES appears to be superior as compare to other models for prediction of recovered cases. At the end, on the bases of these best models, we forecast future 4, July to 14, August 2020, which can help decision making in public health and other sectors for the entire country. Furthermore, this work may help in remembering present socio-economic and psychosocial misery affected by COVID-19 amongst the public in Pakistan.

## Data Availability

We used daily COVID-19 conformed, deaths, and recovered cases for Pakistan. The dataset was obtained by WHO, the each series ranges from 10, March 2020 to 3, July 2020.

https://www.who.int/

